# Evidence for Aerosolized Environmental Bacterial Endotoxin as an Environmental Health Hazard

**DOI:** 10.1101/2025.10.02.25337178

**Authors:** Keziyah Yisrael-Gayle, Edward Helderop, Chris Morin, Daniel Polk, Xinru Qiu, Hovanness Dingilian, Diana del Castillo, Veronica Penuelas, Talyssa M. Topacio, Linton Freund, Ryan Drover, Daniel Gonzalez, Sumaya Troy Alaama, Ahmad Husari, Esther Caroline McGowan, Robin Merwyn, Jon K. Botthoff, Malia Shapiro, Rishi Swaran Kongari, Emma Aronson, Adam Godzik, Jenna LeComte-Hinely, Ran Wei, David Cocker, David D. Lo

## Abstract

The high incidence of asthma near California’s Salton Sea is attributed to dust emitted from the exposed lakebed. A survey of clinical symptoms conducted in the region revealed distinct spatial distribution with strong geospatial correlations between asthma symptoms and prevailing wind direction as well as with PM10 levels. Mice exposed to dust collected across the region showed lung inflammation which was TLR4-dependent; dust inducing the strongest inflammation showed the highest concentrations of LPS. Inflammatory responses and LPS distribution showed a distinct spatial distribution. Environmental halophiles cultured from Salton Sea water produced LPS that induced similar lung inflammation in mice. These results suggest that environmental bacterial LPS entrained in the dust promotes asthma-like disease; thus, perturbations of “natural” ecosystems may produce significant health impacts.

## 1 Introduction

The Salton Sea (SS) is California’s largest lake; the lake continues to retreat due to evaporation and declining freshwater inputs, with increasing areas of exposed lakebed (playa) emitting high levels of dust (1–3). Residents in the region closest to the lake suffer from high levels of asthma that is generally attributed to this playa dust (4,5), though the specific contribution or mechanism of induction of asthma symptoms has not been determined.

## 2 Materials and Methods

### In vivo environmental exposures

8–9-week-old male and female C57BL/6J mice were purchased from Jackson Laboratory. Mice were housed under a 12-hour light/dark cycle with ad libitum access to food and water. After a 1-week acclimation period, mice were housed 2–3 per cage, placed in an environmental aerosol exposure chamber, and exposed to dust from the Salton Sea region for either 48-hours or 7-days.

Upon completion of exposure, mice were euthanized and analyzed for cellular recruitment, gene regulation, and histological alterations. All procedures were in accordance with approved Animal Use Protocols, institutional and NIH guidelines, and experimental design follows the ARRIVE 2.0 guidelines.

### Salton Sea dust collection and extraction

Passive dust collectors were placed at various locations surrounding the sea. These collectors consisted of a round Bundt pan coated with Teflon, lined with Kevlar mesh, and filled with glass marbles, as described in Biddle et al. (6). To recover material, marbles were rinsed with water, and the resulting dust suspension was filtered through sterile 0.2 µm filters, lyophilized, and resuspended for aerosolization in our environmental exposure chambers.

### Bacterial isolation from Salton Sea water

Water samples were collected from seven sites around the Salton Sea using sterile bottles. Samples were filtered through 70–100 µm strainers to remove large debris. Ten microliters of each filtered sample were plated on high-salt agar plates (5% NaCl) and incubated at 37°C. Further colony isolation was performed on agar plates containing both high salt (5% NaCl) and high sulfur (0.4% MgSO₄). Bacterial 16S sequencing and identification of isolated halophiles were conducted by GENEWIZ (Azenta Life Sciences).

### Exposure Chambers

#### Large chamber studies

Exposure studies were conducted using dual-animal chambers as previously described (7,8). Relative humidity, temperature, and atmospheric pressure were monitored, along with particulate matter (PM) size and concentration using a scanning mobility particle sizer (SMPS), to ensure consistent PM exposure. Animals were exposed to either filtered air (control) or aerosolized Salton Sea dust for 48-hours or 7-days.

#### Small chamber studies

Seven Gram-negative halophilic bacterial isolates from Salton Sea water were cultured in high salt (5%) LB media, and pure LPS was extracted from each using a phenol-based extraction and purification kit by Bulldog-Bio. Male and female mice (8–9 weeks old) were exposed to 1mL of this extracted material, LPS from commercial *E. coli,* or Phosphate buffered saline (PBS) for 5 minutes in a modified individual animal chamber equipped with a nebulizer (LPS concentration: 5.91*10^5^ Eu/mL). Mice were allowed to rest overnight before collection of bronchoalveolar lavage fluid (BALF) and lung tissue for flow cytometry and histological analysis.

#### Flow Cytometry

BALF and left lung lobes were collected for flow cytometry analysis. BALF and digested lung tissue were stained for the following surface markers: anti-CD45 FITC, anti-CD19 Percpcy5.5, anti-CD3 AF700, anti-CD4 BV711, anti-CD8 PE-CF594, anti-SiglecF APC, anti-Ly6G BV510, anti-CD11c PE-Cy7, anti-CD11b BV421, and anti-IgE PE. Cells were fixed in 1% PFA and samples were run the following day using the Agilent NovoCyte Quanteon Flow Cytometer. Upon excluding dead cells identified by positive staining with Zombie Yellow dye, cellular populations were identified using the following surface markers: CD45^+^CD11c^+^ SiglecF^+^ for alveolar macrophages, CD45^+^CD11b^+^Ly6G^+^ for neutrophils, CD45^+^CD11b^+^SiglecF^+^ for eosinophils, CD45^+^CD3^+^ for T cells, CD45^+^ CD3^+^ CD4^+^ for CD4 T cells, CD45^+^ CD3^+^ CD8^+^ for CD8 T cells, CD45^+^CD19^+^ for B cells, and CD45^+^CD19^+^ IgE^+^ for IgE B cells. All gating and analysis were done using FlowJo. This data was then transferred into GraphPad Prism 9 for production of graphs and statistical analysis.

Gating was performed using FlowJo. Graphs and statistical analysis were generated using GraphPad Prism 9, with significance set at p < 0.05 using t-tests or ANOVA.

#### Nanostring gene expression profiling

RNA was extracted from flash frozen right lung lobes of exposed and control animals for NanoString analysis. RNA was extracted using a TRIzol based extraction method. Approximately 100 mg of tissue was ground using liquid nitrogen in a mortar and pestle to pulverize the tissue into a fine dust. Following this, tissues were placed in TRIzol and centrifuged for dissociation of RNA. The supernatant was then collected and mixed with Chloroform to allow for further separation before centrifugation. The aqueous phase of the centrifuged product was mixed with isopropyl alcohol and centrifuged before a series of washes with 75% ethanol. RNA was resuspended in DEPC-Treated water and concentration and purity were measured using a Nanodrop 2000 spectrophotometer. For Nanostring analysis, 50 ng of RNA was analyzed using the NanoString nCounter Mouse Immunology Panel to assess gene regulatory changes. Differential expression was calculated and normalized based on housekeeping genes. Positive controls and p-values were adjusted using the Benjamini-Hochberg method.

#### 10x Chromium single cell flex gene expression profiling

After a 7-day exposure to dust material collected from Wister, a site on the southern end of the Salton Sea, BALF was collected and processed for flex gene single cell profiling. Samples were processed using the 10x Genomics Flex kit and downstream analysis was performed in-house using Cell Ranger v8.0.1.

#### 10x Single-Cell RNA-Sequencing Data Processing and Integration

Single-cell RNA-sequencing (scRNA-seq) data were generated using the 10x Genomics Chromium platform and consisted of three libraries: two control replicates (Control_1 and Control_2) and one exposure group (Exposed_3). Raw FASTQ files were processed using Cell Ranger v8.0.1 (10x Genomics), aligned to the mm10-2020-A mouse reference genome using the Chromium Mouse Transcriptome Probe Set v1.0.1. Gene expression matrices were then imported into Python using Scanpy v1.10.3, and each dataset was annotated with batch and condition metadata for downstream integration. Cells expressing fewer than 200 genes or more than 10,000 genes, as well as those with >20% mitochondrial transcripts, were filtered out to remove low-quality cells and potential doublets. Genes expressed in fewer than 3 cells were excluded. Quality control metrics—including gene count, cell count, and mitochondrial content distributions—were visualized to assess data integrity pre- and post-filtering. Quality control and filtering steps were performed using AnnData v0.10.9, NumPy v1.26.4, and Pandas v2.2.3. A computational doublet detection strategy was employed using simulated doublets, PCA-based dimensionality reduction, and k-nearest neighbor density estimation (k=50). Cells identified as putative doublets based on a conservative threshold (mean + 2 SD of simulation scores) were removed. Dimensionality reduction and distance computations were carried out using scikit-learn v1.4.2. Gene expression was normalized to 10,000 total counts per cell followed by log1p transformation. Highly variable genes were selected using the Seurat method. A curated list of macrophage-relevant genes, including Tnfrsf1a, Tnfrsf1b, Marco, and Pparg, was manually retained in the feature set to preserve biologically relevant pathways. Data were z-score scaled, and principal component analysis (PCA) was performed, retaining 50 components. Harmony was used for batch effect correction while preserving biological variation. A k-nearest neighbor graph was constructed using Harmony-corrected embeddings. Leiden clustering was applied on the neighborhood graph with a resolution parameter of 0.8 to identify cell populations. Uniform Manifold Approximation and Projection (UMAP) was used to visualize cellular relationships.

Condition and batch distributions were visualized across the UMAP space to evaluate integration quality. All dimensionality reduction, clustering, and visualization steps were performed using Scanpy v1.10.3 and supporting packages. Wilcoxon rank-sum tests were performed to identify differentially expressed genes (DEGs) across clusters using scanpy.tl.rank_genes_groups. DEGs were filtered for statistical significance (FDR-adjusted p < 0.05) and minimum log fold-change thresholds. Clusters with fewer than two cells were excluded from statistical testing to maintain robustness. Multiple testing correction was applied using the Benjamini-Hochberg procedure. Cell types were assigned based on a curated reference of canonical marker genes for immune, stromal, and structural populations. Marker scores were computed using a weighted strategy that incorporated both the number of marker genes expressed and their expression intensity. Final cell type labels were assigned based on the highest scoring cell identity per cluster. Cells with ambiguous marker expression were designated as “Unknown.” Major annotated populations included classical and non-classical monocytes, macrophage subsets (e.g., alveolar, M1, M2a/b/c), dendritic cells (DC1, DC2, pDC), neutrophils, eosinophils, epithelial and endothelial cells, fibroblasts, CD4+ and CD8+ T cells, NK cells, B cells, and plasma cells.

#### Limulus Amebocyte Lysate Assay

Dust, water, and playa samples collected from around the Salton Sea were tested for endotoxin concentrations using the ToxinSensor Chromogenic LAL Endotoxin Assay Kit (GenScript, ABIN491527). Samples were processed via manufacturer instructions and run neat, as well as at 1:10, 1:50, 1:5000, and 1:10000 dilution concentrations. Absorbance values were measured at a wavelength of 545 nm using a SpectraMax ID5. Concentrations were then back calculated from the standard curve and dilution factors were corrected for to give final Endotoxin concentrations for each sample. Values were then standardized based on the length of time the dust collectors were out for collection of material.

#### Sandwich ELISA assay

LPS was also measured in samples through a competitive ELISA (absorbance 450 nm; ELISA Kit - LS-F15272, LSBio, Seattle, WA), following manufacturer’s specifications, using a SpectraMax iD5 microplate reader (Molecular Devices, San Jose, CA). Samples from Salton Sea collection sites were run neat and in three 1:5 serial dilutions. An additional positive control of lipopolysaccharides from *E. coli* O55:B5 (LPS; L2880) at a concentration of 0.1 ng/mL with three 5-fold serial dilutions was also measured. Endotoxin units were calculated based on a serial fold dilution of an endotoxin standard provided by the manufacturer.

#### Histology

For histological analysis, lungs were inflated with 0.3ml of 10% neutral buffered formalin (NBF). Lungs were then excised and placed in 10% NBF overnight. Lungs were washed in 1x PBS and transferred to 70% ethanol. Tissues were sent to University of Irvine Medical Center for paraffin embedding and sectioning at 5 µm using a microtome. Sectioned were deparaffinized and stained with hematoxylin and eosin (H&E) to analyze cellular recruitment and histological alterations.

#### Community clinical symptom survey

The clinical symptom survey study (Supplementary Figure S1) was approved by the University of California, Riverside Institutional Review Board, protocol HS22-154. HARC and HDR@UCR drafted a survey with 42 questions, which was modeled on a standardized questionnaire from the Global Asthma Network, originating from the International Study of Asthma and Allergies in Childhood (ISAAC) program. Survey questions are available from the authors on request. Results are from an address-based survey of households in eastern Riverside County and northern Imperial County. HARC (via Ace Printing) mailed a “survey package,” which included an invitation letter (in English and Spanish), a paper survey in English, a paper survey in Spanish, a pre-paid return envelope, and a $2 bill as a pre-incentive that the recipient could keep regardless of whether they took the survey. Further, the invitation letter explained that the first 1,000 households to complete and submit the survey would receive a $20 Visa card as compensation. Ace Printing purchased a list of 6,941 households likely to have children and mailed the survey package to these households. In addition, a flyer with a URL link and QR code (to take the survey online) was mailed to an additional 18,000 households (regardless of whether they were likely to have children or not). These flyers likewise offered a $20 Visa card. The mailed instructions asked the adult in the household most familiar with the child to take the survey (the survey thus was often completed by the child’s parent or grandparent). If a household had more than one child, the survey questions concerned the oldest child under the age of 18 in the home. The survey was launched on August 18, 2023, and the survey was closed on October 31, 2023. A total of 840 completed surveys were received. All paper survey results were entered into the online survey platform Sogolytics. The results were downloaded, cleaned, and analyzed using SPSS (Statistical Package for the Social Sciences). *Demographics*: The average age for surveyed children was 12 years old. Among surveyed children, 55.8% were boys, and 44.2% were girls. About 93.8% were Hispanic/Latino. The median household size was 5.0 people.

Nearly half (49.4%) of households had a total annual income of less than $35,000, and a majority (50.9%) of households were at or below the federal poverty level.

#### Geospatial analysis correlating symptom survey results and environmental data

In addition to the surveyed households, environmental and water contaminant data were collected for the study area. Specifically, groundwater contaminant data were sourced from the California Water Boards Groundwater Ambient Monitoring and Assessment Program (GAMA) (9), municipal water contaminant data from the Safe Drinking Water Information System (SDWIS) (10), air quality data from the Environmental Protection Agency (11), and wind direction from the Global Wind Atlas (12). From the municipal water and groundwater data, ten chemicals were used, based on the possible mechanisms of interest: arsenic, iron, aluminum, nitrates, nitrites, phosphates, selenium, uranium, chromium, and manganese.

In our study area, there were six different municipal water suppliers present. Each of these municipalities publishes contaminant data for each of our chemicals of interest at each site monitored. The surveyed households located with the borders of a municipal water supplier (95% of households surveyed) were assigned contaminant data based on the average of the farthest downstream sampling sites for each municipality. The GAMA data, meanwhile, are a composite dataset of sampled locations from at least nine different organizations. The sampling locations were used to construct interpolated surfaces for each contaminant of interest for our study area, and those surfaces were used to assign contaminant values for the five percent of households located outside the boundary of any municipal water supplier. The same interpolation process was used to generate a continuous surface of the atmospheric pollutant PM10, with the resulting values assigned to each household. The predominant wind direction across the Salton Sea is along its long axis, from the northwest to the southeast. Each surveyed household was given a vectorized value that denotes is position relative to the predominant wind direction, as in Figure 1. The relative angle between a vector drawn between each household and the midpoint of the Salton Sea and the predominant wind direction was calculated.

**Fig. 1.**
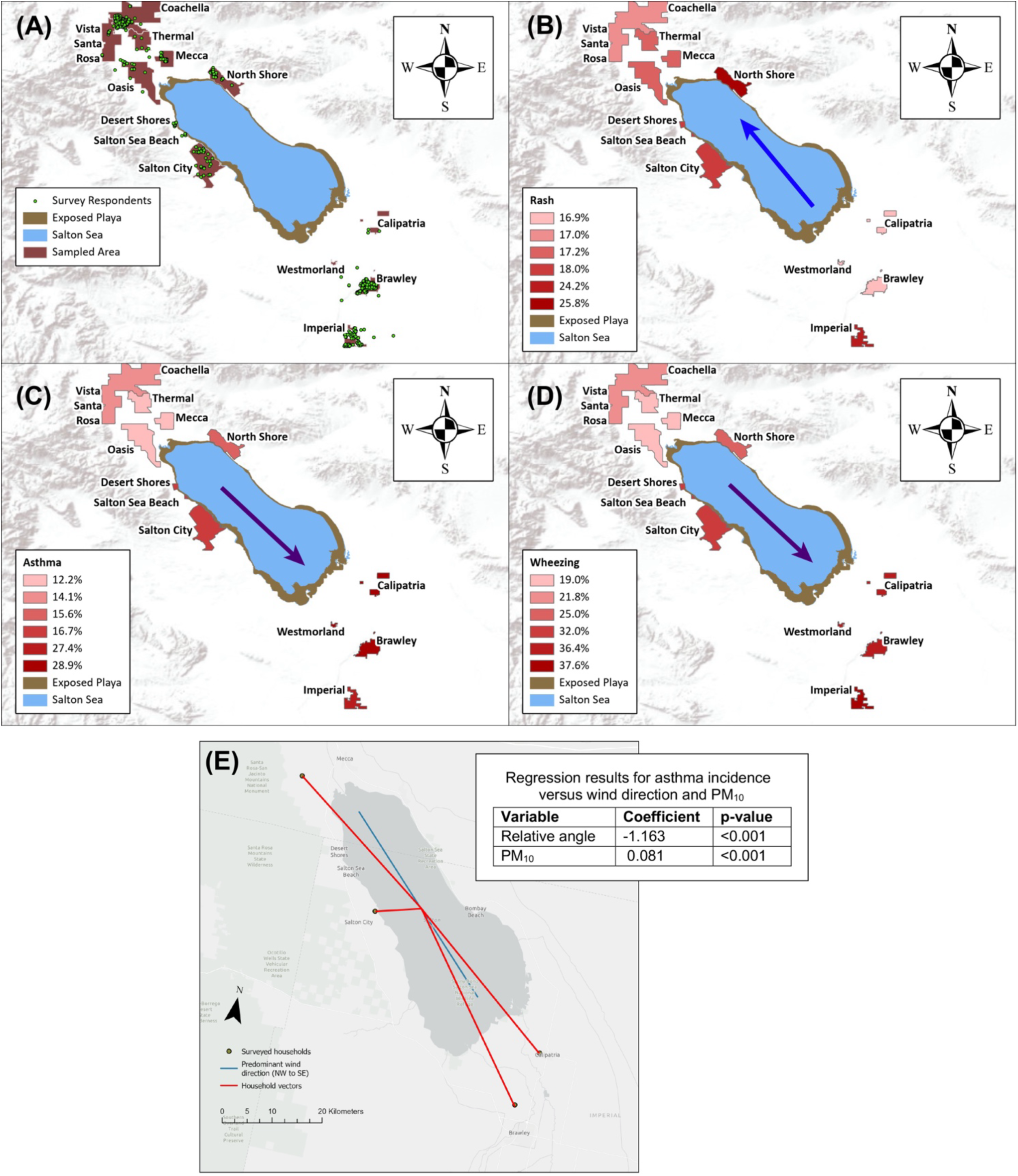
Geographic distribution of symptoms correlating with environmental conditions. Survey responses were grouped into local cohorts for purposes of calculating symptom incidence. Note that the incidence reflects the percentage of responding families with the specific symptom, rather than the total incidence in the population. A, green dots indicate survey respondents; these were mainly found within population centers. B, Incidence of skin rash, showing highest incidence in communities at the North Shore and Imperial, north of the Salton Sea (blue arrow). Response numbers: Brawley, Calipatria, and Westmorland *n* = 266. City of Imperial *n* = 99. Desert Shores, Salton City, and Salton Sea Beach *n* = 50. North Shore *n* = 31. Mecca, Oasis, and Thermal *n* = 99. City of Coachella and Vista Santa Rosa *n* = 271. C,D, Incidence of asthma diagnosis and wheezing, demonstrating a spatial pattern in which lower incidence rates were found in households to the northwest and higher incidence rates in households to the southeast (purple arrow). Response numbers for C, D: Asthma diagnosis - Brawley, Calipatria, and Westmorland *n* = 266. City of Imperial *n* = 95. Desert Shores, Salton City, and Salton Sea Beach *n* = 48. North Shore *n* = 32. Mecca, Oasis, and Thermal *n* = 98. City of Coachella and Vista Santa Rosa *n* = 262; Wheezing - Brawley, Calipatria, and Westmorland *n* = 272. City of Imperial *n* = 101. Desert Shores, Salton City, and Salton Sea Beach *n* = 50. North Shore *n* = 32. Mecca, Oasis, and Thermal *n* = 100. City of Coachella and Vista Santa Rosa *n* = 275. E, Asthma and wind direction. Only a small subset of the surveyed households is displayed for clarity. For each household shown, a vector was drawn between it and the midpoint of the Salton Sea, and the angle between that vector and the predominant wind direction was assigned. In this example, the two households to the southeast of the Salton Sea end up with a low relative angle (<10°), the household to the west has a higher angle, (100°), and the household to the northwest has an even higher angle (170°).

Dummy variables were constructed for asthma and rash incidence for the households, and these were used as dependent variables for the models, while the groundwater/municipal water contaminant, surface water contaminant, air quality, wind direction, and demographic data were used as independent variables. Several models were constructed depending on the mechanism of interest, and all models were spatially lagged regressions – several of the input variables exhibit significant spatial autocorrelation, as did the residuals of a spatially agnostic ordinary least squares model. The resulting models function as exploratory tools, shedding light on the explanatory power of each of the independent variables on asthma and rash incidence rates.

## 3 Results

To search for potential environmental sources contributing to the asthma, we performed a clinical symptom survey to assess the incidence and distribution of specific symptoms across the Salton Sea region. Previous estimates of asthma incidence have been less detailed, with relatively small numbers grouped into relatively large geographic regions such as zip codes or census tracts, which make it more difficult to associate symptoms with proximity to potential sources of disease triggers (13).

From discussions with residents, major concerns included asthma, skin rashes, and nosebleeds, so a questionnaire was developed to query related symptoms along with demographic information. The survey gathered approximately 830 responses from families across the region from Eastern Coachella Valley to Imperial Valley, each response including geospatial information. In this region, the population is predominantly Latino/Hispanic, with relatively low household income and related socioeconomic indicators. To correlate symptom incidence with more detailed geographic information, we aggregated responses into local population cohorts. The incidence of asthma diagnosis, as well as related clinical symptoms such as wheezing, had a distinct geographic distribution within the region (Fig. 1). The percentage of families reporting asthma-related symptoms was 12-19% at the northern end of the Salton Sea region, rising to approximately 37% in communities at the southern end. This spatial pattern was specific to asthma-related symptoms; in contrast, the geographic distribution of skin rash and nosebleeds showed the opposite pattern, with a concentration of skin rash notably increased in communities at the northern end of the region.

This pattern in asthma symptoms suggests a specific distribution of environmental factors contributing to disease incidence. Accordingly, we examined several factors, including features intrinsic to the region’s geochemistry, such as groundwater contaminants, as well as air quality and other environmental pollutants. We quantified the impact of these factors using basic exploratory data analysis by constructing several spatially lagged regressions using individual households as the unit of analysis, using an inverse-distance band weights matrix. Because the air quality and water contaminant sampling sites are located throughout the study area but were not measures at each individual household, an interpolated surface was generated for each using sensitivity analysis to determine the most appropriate kriging parameters. Each household was then assigned its relevant contaminant data based on the value of the interpolated surface at the household location. Two notable exceptions include water contaminants for households that received municipal water (instead of an interpolated surface generated from groundwater sampling sites, we used municipal water system contaminant sampling data) and the relative location of each household to the predominant wind direction in the area. For this last factor, the predominant wind direction was generated using historical wind data and for each household we calculated the relative angle between that household and the midpoint of the wind direction vector we generated (Fig 1E).

In general, the geographic distribution of groundwater and municipal water contaminant data, including heavy metal contaminants such as arsenic and selenium (14) was not a good explainer of asthma incidence rates (not shown). This is reasonable considering that asthma would most likely be induced in this population by inhaled aerosols. In addition, groundwater sources are less likely to be a factor since the majority of households receive municipal water. This results in a lower degree of freedom than might otherwise be expected given the sample size, since nearly 800 of the households share one of only six values for each of the contaminants (one for each of the municipal water suppliers present in the study area).

The levels of PM10 and the relative location of each household to the predominant wind direction were strong predictors of asthma in the population. Both of these variables exhibit moderate collinearity – enough that the exclusion of one variable from the regressions does not significantly impact the R^2^ value of the model. The concentration of PM10 is higher to the south of the Salton Sea than to the north, but not by very much – this may be a function of the relative sparsity and non-uniform distribution of air quality testing stations in our study area. This results in a positive but small coefficient in the regressions. Similarly, wind direction is a consistently strong predictor of asthma. Households that are downwind of the Salton Sea and in the direct path of the predominant wind direction in the area report higher asthma incidence rates. Interestingly, survey participants’ household incomes and race/ethnicity data were not significant predictors of asthma. Although this finding may seem contrary to national health disparity trends, in which asthma is often correlated with lower socioeconomic measures, it is likely because the survey’s population is fairly homogeneous – the great majority of participants were Latino/Hispanic and report a relatively low household income. Fig.1E shows the regression results for these factors – note that due to their collinearity, the displayed results are those of two different spatially lagged regressions (one for each independent variable), and not for a single multiple regression.

To assess the contribution of environmental components that may trigger or drive chronic inflammatory lung disease or asthma, we studied the effects of aerosols generated from materials collected near the Salton Sea using an in-vivo environmental system (6,7). When filtrates of dust samples were used, many of the samples induced significant acute neutrophilic inflammation in exposed mice. The patterns of cell recruitment and gene expression contrasted with the allergic inflammatory response to an aerosolized fungal allergen, suggesting that the driving component in the dust was not allergenic (6).

Exposures to dust samples collected at different sites in the area induced similar patterns of neutrophilic inflammation, but the magnitude of the response varied among samples across the region (Fig. 2A,B). As with asthma symptoms, this variation manifested similarly; while significant inflammatory responses were found in response to samples collected at the northern end, higher responses were induced to samples collected at the southern end of the region, most evident for 7-day exposures (Fig. 2A). Acute neutrophil responses were evident in 48-hour exposures as well (Fig. 2B); in these cases, the highest inflammatory responses were to dust collected near Wister (WI) at the southeastern shore.

**Fig. 2.**
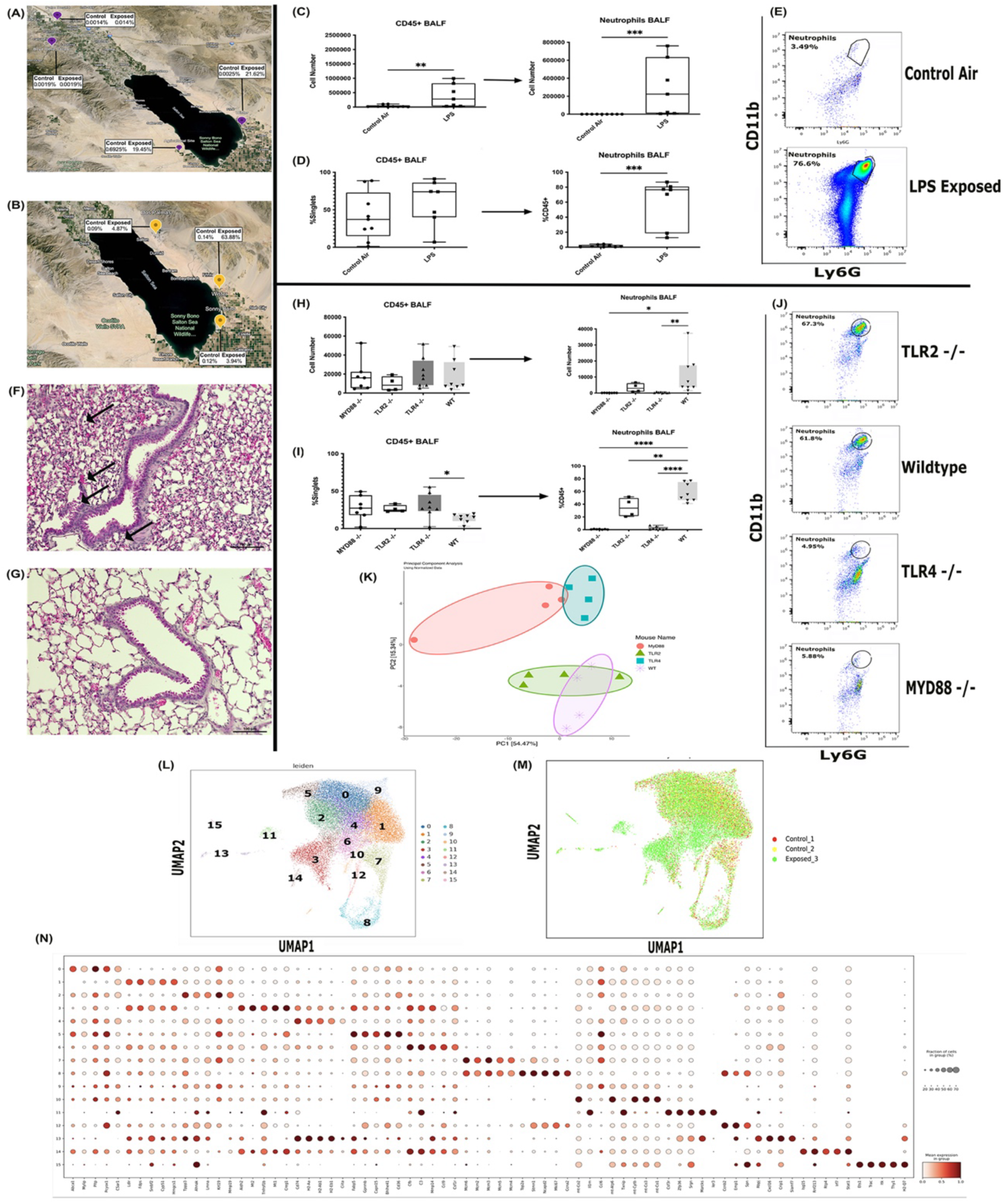
Lung response to Salton Sea dust is TLR4-dependent. **A,B:** Mouse response to Salton Sea dust from various collection sites. Numbers show the mouse inflammatory responses (percentage of neutrophils among CD45+ cells in lavage) to 7-day exposures **(A)** and 48-hour exposures **(B)** in control air and dust exposed mice. **C-E**: Cellular recruitment into the airways after 7-days of continuous exposure to 15 ug/m^3^ of aerosolized commercial *E. coli* LPS **(C)** cell numbers in Bronchoalveolar lavage fluid (BALF); (**D**) cell recruitment shown as percentage of CD45+ BALF cells; (**E**) flow cytometry dot plots showing gating of neutrophils. (P-values: for cell counts, Neutrophils WT vs LPS: 0.0002. Eosinophils WT vs LPS: 0.0079. For cell percentages, Neutrophils WT vs LPS: 0.0003. Eosinophils WT vs LPS: ns.). **F,G**: Histology of neutrophilic recruitment from exposure to Salton Sea dust (AG site dust). (**F**) Lung sections showing infiltration of immune cells and histological alterations in lung tissue (arrows) of wildtype mice after exposure to Salton Sea dust; (**G**) Lung sections showing lack of neutrophil infiltration in lung tissue of TLR4 -/- mice after exposure to Salton Sea dust (scale bars: 100 microns). **H-K**: Response of innate immune receptor knockout mice to Salton Sea dust. (**H**) Bronchoalveolar lavage fluid cell numbers recovered from wildtype (WT), and MyD88, TLR2 and TLR4 knockout mice exposed to Salton Sea dust (AG sample site); **(I)** cell percentages of CD45+ cells and neutrophils from BALF; (**J**) Flow cytometry dot plots showing neutrophils; **(K)** Principal Component Analysis (PCA) of gene expression profiling of lung tissue showing colocalization of WT and TLR2 knockout profiles, distinct from the profiles of TLR4 and MyD88 knockout mice. (P-values. For cell counts, Neutrophils WT vs TLR2ko: 0.1588; vs TLR4ko: 0.0091; vs MyD88ko: 0.0102. Eosinophils WT vs TLR2ko: ns; vs TLR4ko: 0.0164; vs MyD88ko: 0.0136. For cell percentages, Neutrophils WT vs TLR2ko: 0.0024; vs TLR4ko: <0.0001; vs MyD88ko: <0.0001. Eosinophils WT vs TLR2ko: 0.0110; vs TLR4ko: <0.0001; vs MyD88ko: <0.0001.). **L-N:** 10x Single cell sequencing of lung lavage cells after 7-day Salton Sea dust exposure (WI site dust) also confirm patterns of gene expression associated with innate immune activation. **(L)** Numbered UMAP of all identified cell populations in control and WI dust exposed mice; **(M)** UMAP of all identified cell populations by condition (control cells-yellow and red overlayed on exposed cells-green); **(N)** Heat map dot plot highlighting top genes for each cell cluster.

It is notable that the lung inflammatory response to Salton Sea dust showed characteristics of an innate immune response, including an early peak with declining cell recruitment after one week of exposure and gene expression patterns similar to responses to aerosolized innate ligands (6). We examined the possibility that bacterial Gram-negative endotoxin, or Lipopolysaccharide (LPS), is the likely inflammatory ligand in the dust; Figure 2C-E shows a reference response to a commercial preparation of LPS from *E. coli*. Mice were exposed to aerosolized LPS similar to the protocol previously reported (6), and responses included significant neutrophil recruitment. Some eosinophil recruitment was detected, but based on cellular recruitment and gene expression patterns, this was clearly distinct from conventional allergic inflammatory responses to a fungal allergen.

To confirm that the response to Salton Sea dust is indeed a response to LPS, we measured the response of mice lacking receptors for LPS (TLR4 knockout mice), and the receptor for cell wall peptidoglycan and related components of Gram-positive bacteria (TLR2 knockout). We also included mice lacking the innate adaptor protein MyD88 to test the overall requirement for innate immune signaling, as distinct from adaptive immunity to dust components (e.g., allergic inflammation). Mice with genetic knockouts for the innate receptors TLR4, TLR2, and MyD88 were exposed to a Salton Sea dust sample known to induce a strong inflammatory response (AG) for 48-hours. Wildtype and TLR2 knockout mice showed similar strong lung inflammatory responses, while TLR4 and MyD88 knockouts had nearly absent cellular recruitment (Figures 2F,G showing histology of lung infiltrates; 2H-J showing flow cytometry of recruited cells). The pattern of gene expression shown in the Principal Component Analysis (PCA) (Fig. 2K) showed that both TLR4 and MyD88 knockout mice showed dramatically distinct responses, while the TLR2 knockout response co-localized with wildtype mice.

Single cell sequencing of recruited cells in lung lavage from 7-day WI dust exposed mice (Fig 2L, M) confirm activation of pulmonary macrophages (clusters 3, 6, 9, 12, and 14, identified based on expression of F4/80, CAR4, IL1-β, NOS2, CCR1, and MARCO), and induction of genes (Fig 2N) associated with innate immune triggering consistent with neutrophil recruitment (cluster 11, determined by CSF3R, CD11b, and CXCR2 expression) and LPS-induced activated macrophages (cluster 9 and 14, defined by expression of MARCO, NOS2, IL1RN, IL-6, and GBP2/5).

Supplemental Table 1 summarizes gene markers used to define these clusters and their phenotypes. Given this data, the inflammatory response to Salton Sea dust is primarily an innate immune neutrophilic response to LPS. In affected communities, this innate response might be conflated with clinical symptoms of atopic (allergic) asthma.

If LPS were the main component driving lung inflammation in our mouse studies and asthma symptoms in the community, then LPS might be detectable in significant concentrations in dust across the geographic range. We found that LPS concentrations measured in the dust and water samples across the region were detected with the highest concentrations in samples collected near the Salton Sea, both at the northern and southern ends of the lake (Fig. 3A-C), with the highest concentrations by far found in both dust and water samples at the southeastern end, near Wister.

**Fig. 3.**
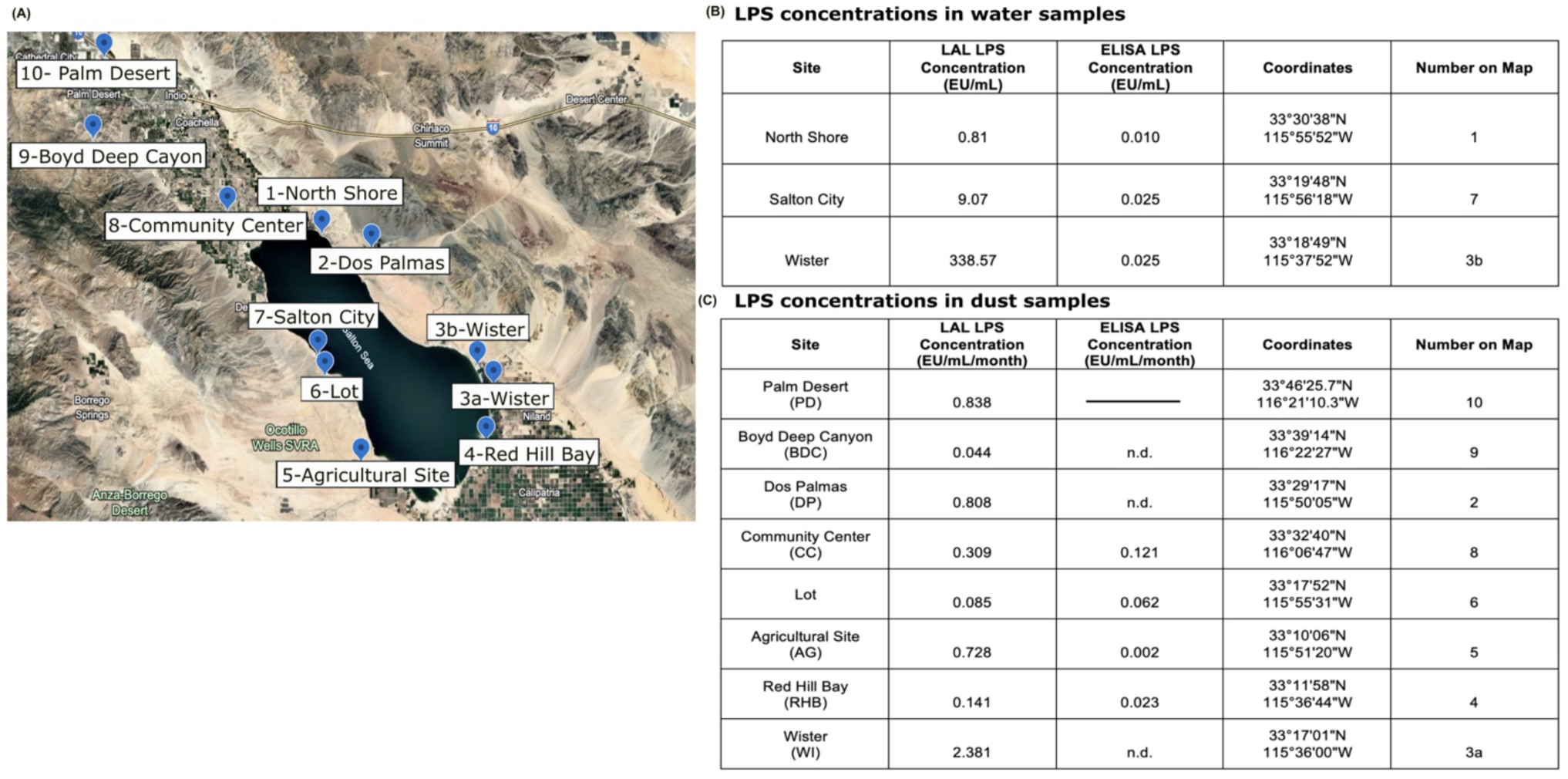
LPS concentrations in dust and water samples from the Salton Sea region. (A) map showing collection sites; (B) LPS concentrations measured in water samples collected from the shoreline of Salton Sea, using LAL and ELISA assays; (C) LPS concentrations measured in dust samples, using LAL and ELISA assays. LAL assays produced a higher signal than ELISA assays, with the ELISA often showing undetectable signals.

The ability to measure the LPS in Salton Sea samples appears to be related to its biological origin. Endotoxin can be present in environments where untreated sewage is dumped into the water resulting in high levels of coliform bacterial LPS in the water (15). In the Salton Sea, LPS may have environmental as well as anthropogenic sources; thus, in testing for LPS, we used two different assays, one using a biological assay based on the limulus amebocyte lysate (LAL), and a competitive ELISA assay, which is a structural assay specific to clinical pathogen (e.g., coliform) LPS. While the biological LAL assay was able to detect LPS in all of our samples, the ELISA was unable to detect signal in most of the same samples (Fig. 3B,C). These results suggest that the main inflammatory LPS found in our dust samples was not from sewage wastewater, and more likely produced by environmental bacteria, such as halophiles in the saline Salton Sea.

To confirm this connection, seven Gram-negative halophile isolates from Salton Sea water samples were cultured in high-salt, high-sulfur media, and LPS was extracted. We then tested whether LPS from these Salton Sea microbes could induce inflammation in mice. Notably, when wildtype mice were exposed to aerosols of LPS extracted from the seven Salton Sea halophilic bacteria, we observed a potent induction of neutrophilic lung inflammation (Fig 4A-C), confirming the inflammatory toxicity of environmental microbial material.

**Fig. 4.**
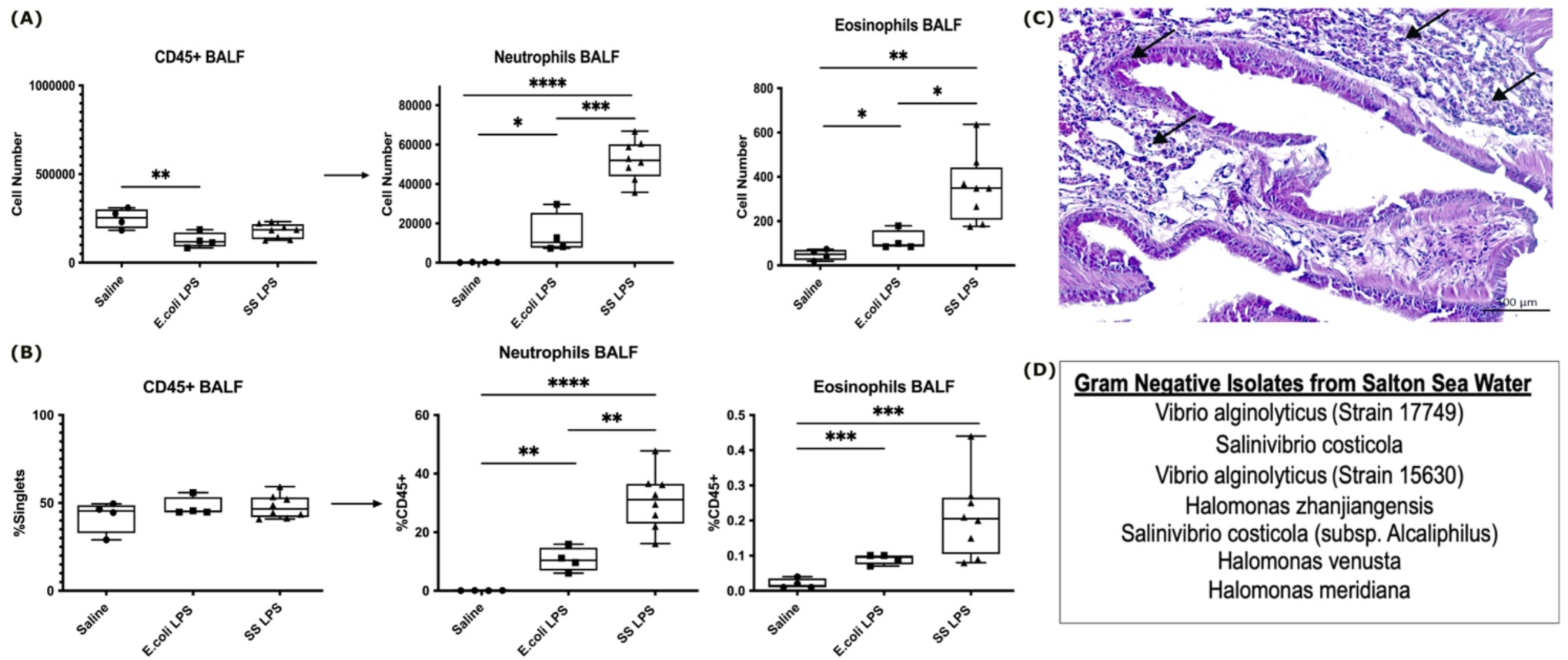
Environmental microbes LPS extraction and mouse lung inflammatory response. (A,B). Recruitment of significant numbers of neutrophils in the lung Bronchoalveolar Lavage Fluid (BALF) but minimal numbers of eosinophils after aerosol exposure to extracted Salton Sea environmental bacteria LPS. (C) histology showing tissue recruitment of neutrophils (arrows).

## 4 Discussion

These studies extend previous studies suggesting that ambient dusts collected near the Salton Sea can induce strong neutrophilic lung inflammation, confirming that this response is an innate TLR4-dependent response to aerosolized LPS. Moreover, the geographic distribution of asthma symptoms among residents in the region correlates strongly with the distribution of LPS concentrations in the ambient dust, driven by the prevailing winds across the Salton Sea. Environmental halophilic bacteria are the most likely source of this LPS, possibly entrained in dusts generated in sea spray and the exposed playa of the drying lake. A recent report showed that the Salton Sea has significant nutrient flux due to agricultural runoff, which contributes to strong microbial growth and accompanying eutrophication with release of decaying microbial components (16). The highest nutrient flux was at the extreme southern end of the lake, where we also measured the highest concentrations of LPS in both water and dust. Collectively, our data suggest a scenario whereby nutrient-driven overgrowth of environmental halophilic bacteria contribute inflammatory LPS for entrainment in ambient dusts.

This dust is carried on prevailing winds into nearby communities where they induce a chronic innate pulmonary inflammation, leading to the high incidence of asthma-like symptoms. Considerations of this scenario should help drive strategies for more appropriate diagnosis and treatment of pulmonary disease in this region, as well as raise questions about a possible role for aerosolized LPS in asthma-like symptoms in other settings

## 6 Author Contributions

Conceptualization: KYG, EH, JL-H, AG, DDL

Methodology: KYG, EH, AH, XQ, ECM, LJ-H, RW, VP, EA, DC, DDL

Investigation: KYG, EH, CM, DP, XQ, HD, DdC, TT, LF, RD, DG, VP, STA, RM, JKB, MS, RSK

Visualization: EH, CM, XQ, DDL, KYG Funding acquisition: DDL

Project administration: DDL

Supervision: DDL, RW, JL-H, EA, AG, DC Writing – original draft: KYG, EH, DDL Writing – review & editing: KYG, EH, DDL

## Data Availability

All data produced in the present study are available upon reasonable request to the authors

## 8 Supplementary Material

**Fig. S1.**
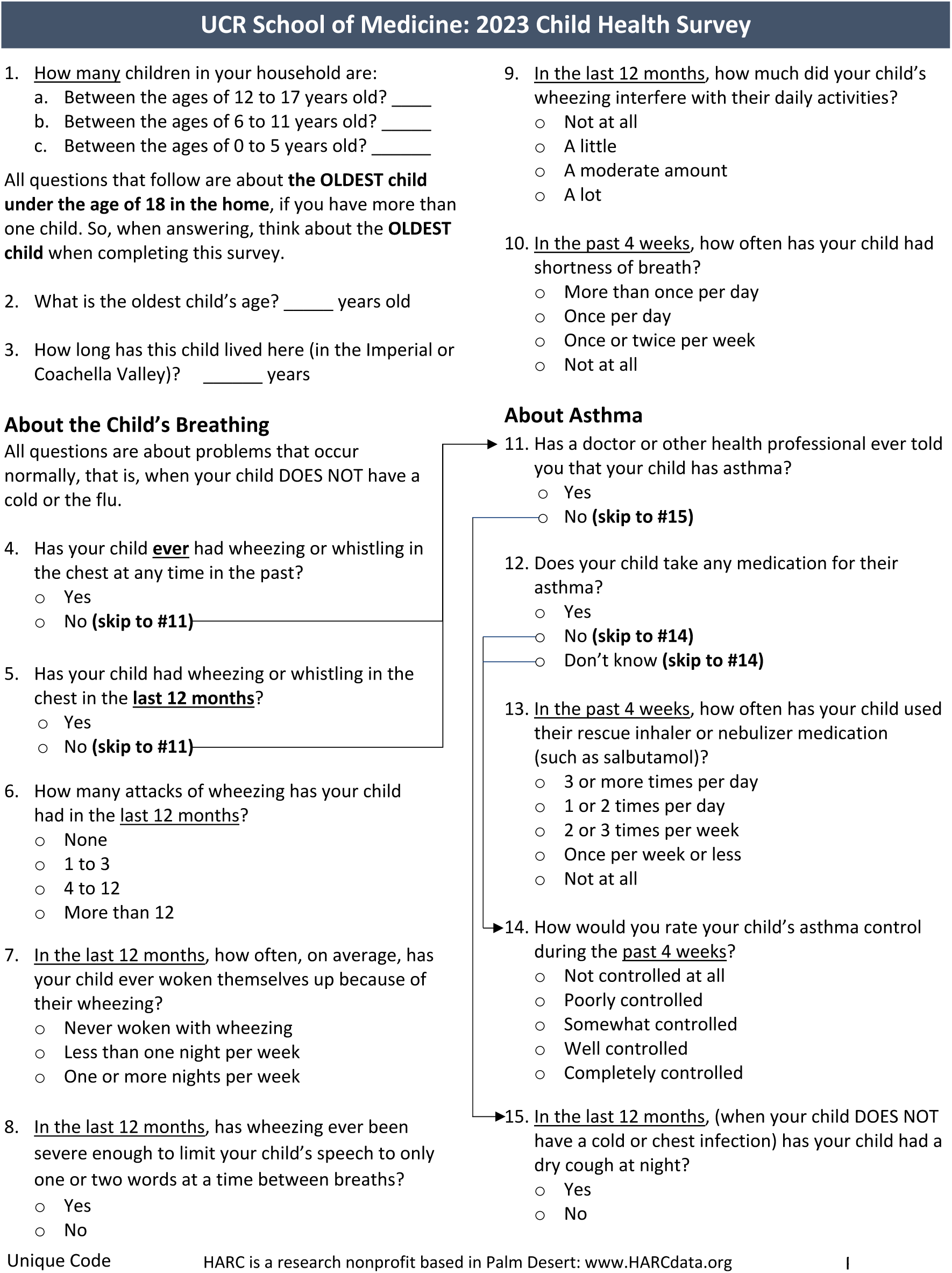
Survey questions. See Materials and Methods, <u>Community clinical symptom survey</u> for details.

---

**Table S1.**
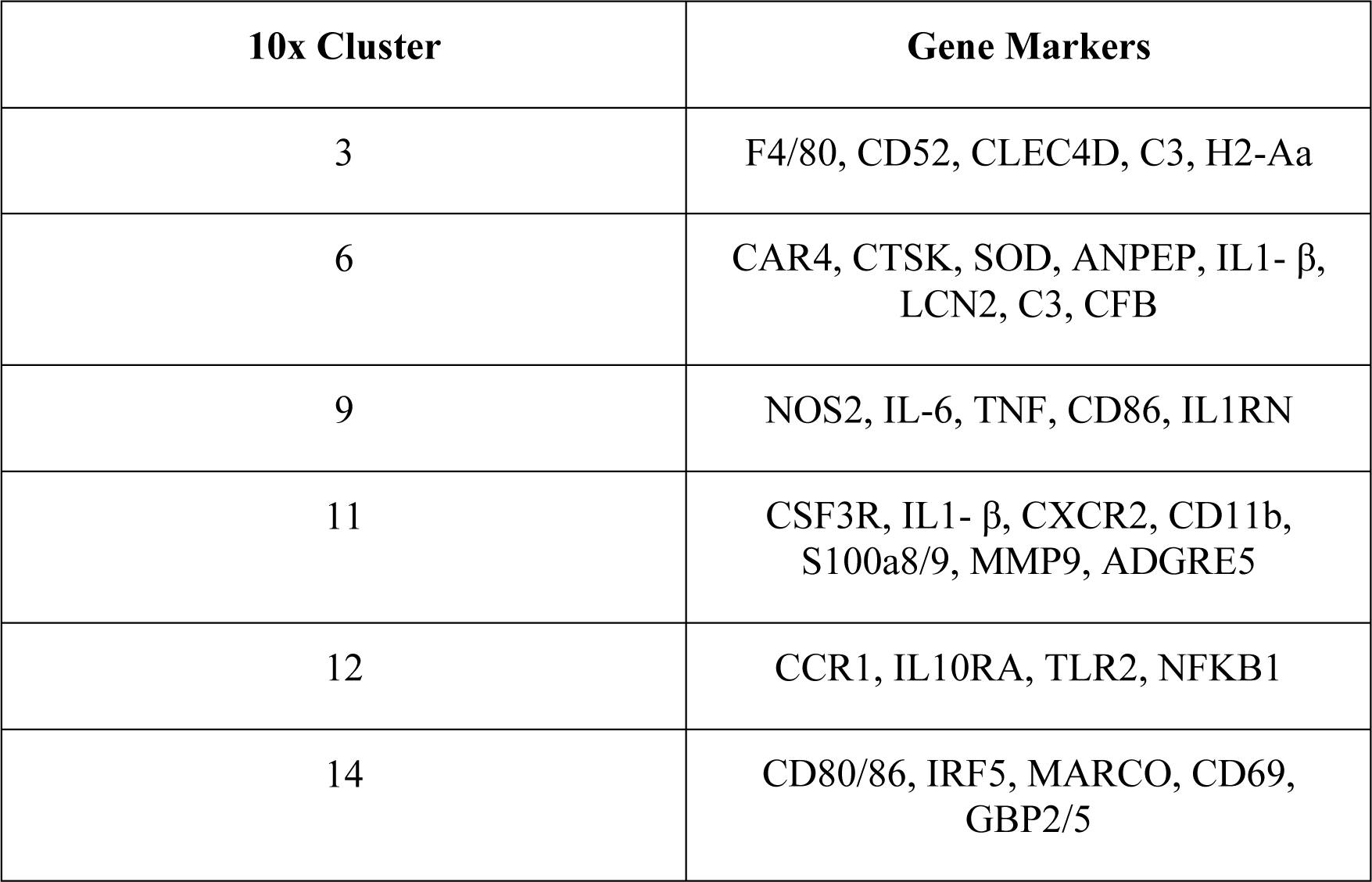
Cluster-Defining Genes Identified by 10x Single-Cell Analysis. Cell-specific gene markers from cell clusters identified in Fig. 2L and N are listed here. See Materials and Methods, <u>10x Single-Cell</u> <u>RNA-Sequencing Data Processing and Integration</u> for details.

| 10x Cluster | Gene Markers |
| --- | --- |
| 3 | F4/80, CD52, CLEC4D, C3, H2-Aa |
| 6 | CAR4, CTSK, SOD, ANPEP, IL1- $\beta$,<br>LCN2, C3, CFB |
| 9 | NOS2, IL-6, TNF, CD86, IL1RN |
| 11 | CSF3R, IL1- $\beta$, CXCR2, CD11b,<br>S100a8/9, MMP9, ADGRE5 |
| 12 | CCR1, IL10RA, TLR2, NFKB1 |
| 14 | CD80/86, IRF5, MARCO, CD69,<br>GBP2/5 |

